# A Change of Heart: The Effect of High Flow Arteriovenous Fistulas on Cardiovascular Outcomes - a Systematic Review and Synthesis without Meta-analysis (SWiM)

**DOI:** 10.1101/2025.02.26.25322822

**Authors:** NA Shah, ME Hind, ZH Endre, BJ Cochran, TJ Barber, JH Erlich

## Abstract

**Background:** Although arteriovenous fistulas (AVF) are widely accepted as the gold standard form of dialysis access for hemodialysis patients, high AVF flow may impose significant hemodynamic stress, potentially contributing to adverse cardiovascular events, including heart failure (HF). Understanding this relationship is crucial for improving patient care.

**Aim:** To describe the relationship between AVF flow and cardiac outcomes.

**Methods:** A comprehensive search of the PubMed, EMBASE, and Cochrane databases was used to identify observational studies and randomized controlled trials reporting an effect of AVF flow on cardiac outcomes (clinical, echocardiographic, and biomarker). Due to study heterogeneity, meta-analysis was not feasible. Synthesis without meta-analysis (SWiM) was performed using vote counting of direction of effect as the primary outcome.

**Results:** Higher AVF flow rates were consistently associated with increased incidence of HF and worsening HF symptoms. Cardiac imaging revealed left ventricular dilation and reduced left ventricular ejection fraction in patients with high-flow AVFs. Elevated biomarkers, such as natriuretic peptides, corroborated the adverse cardiovascular effects of high AVF flow.

**Conclusions:** This systematic review and synthesis without meta-analysis showed a positive relationship between AVF flow and clinical, echocardiographic, and biomarker cardiovascular outcomes, but an inverse relationship between AVF flow and mortality. The methodological heterogeneity of studies highlights the need for well-designed prospective research with standardised definitions of high flow AVFs and measures for reporting of cardiovascular outcomes.

## Introduction

Chronic kidney disease (CKD) is associated with a markedly increased risk cardiovascular (CV) disease, increased mortality, diminished quality of life, and poorer health outcomes across all domains of health compared to individuals with normal kidney function ^1,2^. The overlap between traditional risk factors for heart disease and those for kidney disease underlies the frequent coexistence of these conditions. A high incidence of atherosclerotic CV disease even in the early stages of CKD is well documented ^3^. CV mortality remains significantly elevated in CKD even after adjusting for established CV risk factors, confirming CKD as an independent risk factor for CV death ^2,4^. Even mild renal dysfunction, such as isolated albuminuria, is strongly predictive of adverse CV outcomes ^5-8^.

This elevated risk is attributable to both traditional and CKD-specific risk factors including albuminuria, sympathetic overactivation, disordered calcium-phosphate metabolism, anemia, extracellular fluid overload, electrolyte imbalances, oxidative stress, systemic inflammation, malnutrition, uremic toxin accumulation, and endothelial dysfunction. Consequently, patients with CKD face heightened risks of atherosclerotic complications, arrhythmias, and heart failure (HF), which increases with severity of CKD ^9^, with CV disease accounting for up to 50% of deaths in patients with ESKD on dialysis ^10,11^. This elevated risk is attributable to both traditional and CKD-specific risk factors including albuminuria, sympathetic overactivation, disordered calcium-phosphate metabolism, anemia, extracellular fluid overload, electrolyte imbalances, oxidative stress, systemic inflammation, malnutrition, and endothelial dysfunction. Consequently, patients with CKD face heightened risks of atherosclerotic complications, arrhythmias, and heart failure (HF), with CV disease accounting for up to 50% of deaths in patients with end-stage kidney disease (ESKD) on dialysis ^10,11^.

In ESKD, adequate vascular access is essential, and arteriovenous fistulas (AVFs) are the preferred modality ^12^, t^12^. Though they are not without complications. When a high-pressure, high flow artery is connected to the low-pressure venous system, there are immediate effects on the systemic circulation, followed by chronic hemodynamic changes if the shunt is left uncorrected. After AVF creation, mean systemic arterial pressure, effective systemic blood flow, total and pulmonary peripheral resistances decrease, whilst heart rate, cardiac output, stroke volume, left atrial pressure, and pulmonary arterial pressure increase ^13^. Despite the variety of changes to multiple haemodynamic measures, there is no significant change in blood pressure ^14^. The immediate reduction in total peripheral resistance causes an increase in venous return and a subsequent rise in cardiac output driven by an increase in myocardial contractility and heart rate ^14^. Given the requirement for permanent vascular access, the enduring haemodynamic changes cause lasting structural and functional changes to the heart. Despite improvements in afterload once on regular haemodialysis, AVFs result in the development of left ventricular hypertrophy, right ventricle remodelling and increases in left atrial volumes ^15^. Studies have noted up to a 50% increase in the incidence of left ventricular hypertrophy 3 months after fistula creation surgery ^16^. These structural changes are proportional to the magnitude of AVF flow, with onset within weeks of AVF creation ^17^. Cardiac biomarkers associated with an increased mortality in HD patients, such as interleukin-6 (IL-6), N-terminal pro-brain natriuretic peptide (NT-proBNP), and troponin (cTnI), have also been shown to rise following AVF creation ^18,19^. In patients who undergo flow reduction procedures, serum measurements of NT-proBNP and mid-regional pro-atrial natriuretic peptide decrease by up to 67% and 25%, respectively ^20^. Decreases in left ventricle mass, improvements in measures of diastolic dysfunction, and an almost 50% improvement in the rate of pulmonary hypertension have also been described following reductions in AVF flow ^21^. Left ventricular hypertrophy has been shown to partially regress once AVFs are made non-functional, and resolution of symptoms can be seen in patients with high flow AVFs following flow reduction surgery ^22,23^.

Though case reports and some observational studies have associated high flow AVFs with the development of heart failure, studies looking at hard outcomes such as incident heart failure or death have had mixed results leaving the exact effect of high flow AVFs on cardiac function to remain poorly understood. This study systematically summarised the body of evidence examining the impact of AVF flow on incident heart failure, death, cardiac imaging and cardiac biomarkers.

## Methods

### Data Sources and Search Strategy

An initial search strategy was registered with PROSPERO (CRD42023457290) and run in June 2022 searching across 3 electronic databases (PubMed, EMBASE, and Cochrane Central Register of Controlled Trials) without language restriction. The search was re-run on February 12, 2024. The search strategy was developed to identify published reports of clinical trials and observational studies examining the effects of AVF flow on CV outcomes. Index terms, subject sub-headings, and some word truncations related to the intervention, study design, and outcome measures, according to each database, were used to map all possible key words (Supplementary Table 1).

### Participants/population

Study inclusion criteria were: 1) Adult population (>18 years); 2) The presence of a dialysis access AVF or AVG at the time of the study. Exclusion criteria included: 1) patients on continuous renal replacement therapy, 2) patients on peritoneal dialysis, 3) patients on haemodialysis using a vascular access catheter, without an AVF/AVG. Randomised control studies and prospective/retrospective observational studies were included.

### Outcome(s)

The primary outcome was incident heart failure as defined by each individual study. Key secondary outcomes included measurements of general cardiac performance (cardiac output or cardiac index), cardiac parameters on transthoracic echocardiogram (TTE), and biochemical markers associated with cardiac function. Each study assessing general cardiac performance used one of the following methods: right heart catheterization, cardiac magnetic resonance imaging, TTE, thermodilution, chemical indicator dilution, pulse wave analysis, bioimpedance, and ballistocardiography. TTE parameters were categorized into four groups: structural changes, systolic function, diastolic function, and pulmonary hypertension. Biomarkers were categorised as either related to inflammatory state, or volume status (Supplementary Table 2).

### Data Extraction

The search strategy was defined by all members of the review team. After duplicate records were removed, all remaining articles were independently assessed by 2 reviewers (NS/MH) for eligibility based on the pre-defined inclusion and exclusion criteria. References deemed relevant by either reviewer during initial screening of titles and abstracts were included for full text article review (NS/MH). Any discrepancies were adjudicated by a third researcher (BC). Article selection was documented using the PRISMA ^24^ study flow chart quantifying the number of articles reviewed, duplicate results, and articles eventually included. From each included study, reviewers (NS/MH) extracted the following information: study information - author information, year published, and publication journal; participant data - number of participants, study comparator groups, outcome, inclusion and exclusion criteria; study outcomes - as defined above; potential study biases.

### Risk of Bias/Quality

Risk of bias (ROB) was assessed for all included RCTs using the revised Cochrane risk-of-bias tool for randomised trials (RoB 2) ^25^ and for observational studies using the Newcastle-Ottawa Scale ^26^. Two independent reviewers (NS/MH) performed ROB assessments. Discrepancies were resolved by a third researcher (BC). The Grading of Recommendations Assessment, Development, and Evaluation (GRADE) approach was used to assess the overall quality of evidence for the primary outcome.

### Strategy for Data Synthesis and Synthesis Without Meta-Analysis

Data was extracted from individual studies to describe key findings. Data was organised based on comparators (measures of AVF blood flow as described by individual studies) and measured outcomes with a focus on incident heart failure. Visual inspection of data suggested meta-analysis would not possible given heterogeneity in study design, comparator groups, and methods of reporting outcomes. The SWiM (Synthesis Without Meta-analysis) method was used as it provides a structured approach for synthesizing quantitative data in systematic reviews when meta-analysis is not feasible ^27^. The method involved nine key steps: 1) Clear justification for grouping of studies based on relevant characteristics, 2) Standardization of metrics for each outcome, 3) Details of alternative synthesis method employed, 4) Study prioritization criteria, 5) heterogeneity investigation, 6) certainty of findings assessment, 7) visualisation of data which clearly link studies to synthesis findings, 8) Synthesis of findings reported in relation to the review question, including contributing studies and certainty assessments, 9) Limitations of the synthesis method and critically evaluated data grouping .

## Results

The initial search strategy identified 4309 publications. After screening, 107 studies with a total of 490,660 participants remained that assessed the relationship between AVF flow and primary or secondary outcomes (median number of patients per study = 49, IQR 78.5) (Figure 1). Many studies reported more than one category of outcome. Of the 37 studies reporting clinical HF outcomes, 36 were observational studies and 1 was a randomised controlled trial: 19 showed a high risk of bias, 3 a moderate risk of bias, and 15 a low risk of bias (Supplementary Table 3 and Supplementary Table 4). Due to heterogeneity in study design, variability in outcome definitions, and inconsistency of statistical analysis between studies, meta-analysis was not feasible. To combine studies, the SWiM guidelines ^27^ with vote counting of direction of effect was employed.

**Figure 1:**
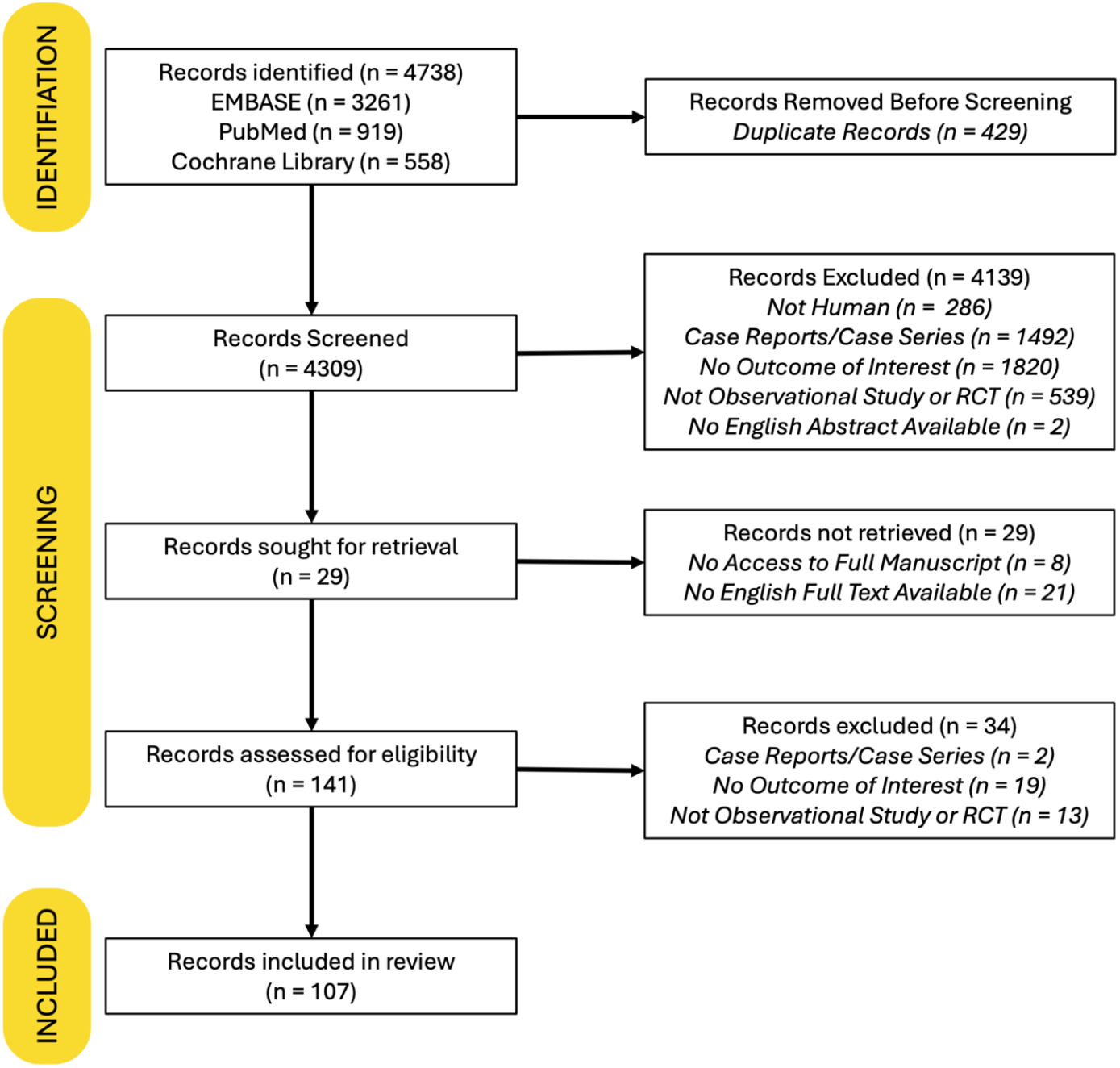
PRISMA Flow Diagram.

Six studies including a total of 2505 participants (median number of patients per study = 137, IQR 423) assessed the primary outcome of incident HF. Four studies identified a positive association between increased AVF flow and incident HF, 1 noted a neutral effect, and 1 a negative association. Two of these studies had a high risk of bias, while 4 had a low risk of bias. Using a composite clinical outcome of prevalent HF and changes in HF symptom burden, 7 studies reported a positive association with AVF flow, while 5 studies found a neutral effect, and 1 study noted a negative effect. Twelve studies evaluated changes in HF symptoms following AVF flow reduction or ligation procedures with all 12 studies reporting symptom improvement after flow reduction. Only one study investigating incident and prevalent HF identified a negative association with AVF flow comparing patients with functional AVFs to those with maturation failure at 8 weeks. Nine studies examined the relationship between AVF flow and all-cause mortality. Among them, 4 studies reported a positive association of mortality with increased AVF flow, 2 found no association, and 3 observed a negative association (Figure 2).

**Figure 2:**
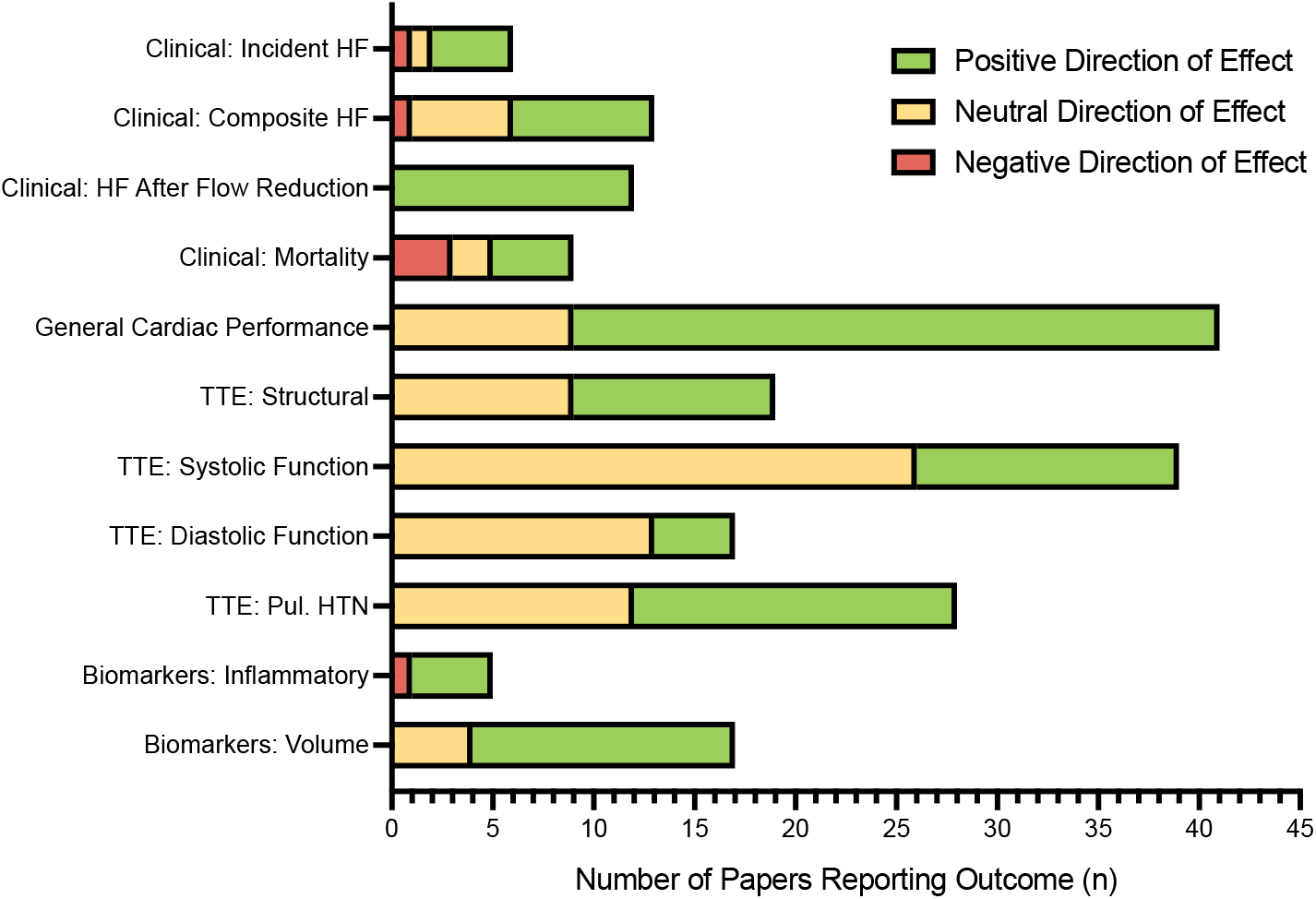
Vote Counting of Direction of Effect for Relationship Between AVF Flow and Cardiovascular Parameters. HF = heart failure; TTE = transthoracic echocardiogram; Pul. HTN = pulmonary hypertension.

Forty-one studies evaluated general cardiac performance parameters using various techniques, as discussed previously. Among these, 32 studies identified a clear association with AVF flow, while 9 reported a neutral effect. In total, 77 studies were included in the analysis of secondary outcomes related to TTE imaging and biomarkers. The 42 studies evaluating TTE parameters were categorized into 4 groups: structural changes, systolic function, diastolic function, and pulmonary hypertension (Supplementary Table 2). Across the TTE categories, studies assessing systolic and diastolic heart function predominantly reported neutral findings with fewer demonstrating a clear association with AVF flow. In contrast, studies evaluating structural cardiac changes and pulmonary hypertension showed a more balanced distribution, with similar proportions reporting either a positive association with AVF flow or no significant effect (Figure 2).

Twenty-two studies evaluated biochemical markers, reporting 17 outcomes focused on volume status and 5 on inflammatory state. Among studies assessing biomarkers of volume status, 13 reported a positive association with AVF flow, while 4 noted a neutral effect. Four studies examining inflammatory markers identified a positive relationship with AVF flow.

Notably, only 1 study investigating TTE or biomarker outcomes identified a negative association with AVF flow (Figure 2).

## Discussion

This study represents the first systematic review of the complex relationship between AVF flow and the spectrum of cardiovascular outcomes. Our findings indicate that higher AVF flows are associated with an increased incidence of HF and worsening of HF symptoms. This observation strongly supports the understanding that the hemodynamic burden of AVFs plays a critical role in driving maladaptive cardiac remodeling and contributes to adverse cardiovascular outcomes in hemodialysis patients. The key secondary outcomes in this study, particularly those related to cardiac imaging parameters and biomarkers, provide robust evidence reinforcing this clinical association.

The compensatory hemodynamic adaptations following the decrease in systemic vascular resistance after AVF formation necessitate an increase in stroke volume to maintain circulatory homeostasis ^28^. This hemodynamic response serves as a key driver of cardiac remodelling, characterized by increases in left ventricular mass/left ventricular mass index, left ventricular end-diastolic diameter/end diastolic volume, and left atrial volume ^29,30^. These structural changes were reflected in the current analysis, where the majority of studies assessing general cardiac performance parameters, such as CO or CI, reported a positive association with AVF flow. Biomarkers of volume status and inflammation were consistent with these findings, with elevated levels of ANP, NT-proBNP, hsCRP, and IL-6 correlating with increased AVF flow. These biochemical markers suggest cardiac stress and endothelial dysfunction may be attributable to mechanical overload and low, oscillatory, or turbulent blood flow patterns.

While AVF flow demonstrated a clear positive association with heart failure, the relationship with all-cause mortality remains uncertain. Three of 7 studies reported an inverse relationship, potentially reflecting the risks associated with low-flow AVFs, including failure to mature, inadequate dialysis efficiency, and underlying peripheral arterial disease or thrombosis ^31,32^. This suggests a U-shaped relationship between AVF flow and mortality which may be attributed, in part, to the haemodynamic burden of higher AVF flow coupled with endothelial dysfunction induced by AVF flow driving elevated levels of pro-inflammatory cytokines, such as IL-6 ^33^. Such a relationship supports the concept of an optimal intermediate AVF flow range associated with reduced rates of cardiovascular events and mortality, in contrast to both low and high AVF flow rates. Although current KDOQI guidelines recommend routine surveillance of high flow AVFs at 6-to 12-month intervals, specific recommendations on the timing and criteria for flow reduction interventions are limited^34^. Emerging evidence, including studies on prophylactic banding of high flow AVFs in post-kidney transplant patients^35^, along with findings from the studies investigating the impact of AVF flow reduction or ligation included in this review consistently demonstrated an improvement in HF symptoms. This suggests that early intervention may play a critical role in mitigating the risk of HF, reinforcing the need for clearly defined guidelines that include upper thresholds of AVF flow.

Across the TTE categories, studies assessing systolic and diastolic function predominantly reported neutral findings, with fewer demonstrating a clear association with AVF flow. In contrast, studies evaluating structural cardiac changes and pulmonary hypertension exhibited a more balanced distribution, with comparable proportions reporting either a positive association with AVF flow or no significant effect (Figure 2). This is particularly noteworthy given the increased risk of heart failure with preserved ejection fraction in patients with CKD ^36^. In patients with heart failure with preserved ejection fraction, where ventricular stiffness and impaired relaxation already compromise diastolic filling, this added volume load should theoretically exacerbate diastolic dysfunction markers. Moreover, ventriculoarterial uncoupling, characteristic of heart failure with preserved ejection fraction ^37^, should be further worsened by the chronic high-output state induced by an AVF, leading to increased LV filling pressures as suggested by elevated E/e’ ratios. The lack of a stronger observed correlation may reflect the significant heterogeneity in study design, outcome definitions, and statistical analysis.

A notable strength of this study is the application of the SWiM guidelines, which enabled the comprehensive synthesis of the existing body of evidence. However, there are several key limitations. One significant issue is the lack of a universally accepted definition of a high-flow AVF, as evidenced by the variability in how different studies defined high-flow status. The KDOQI guidelines suggest a high flow definition of a flow rate > 1-1.5 L/min or >20% of cardiac output, while other studies employ a set threshold of >2 L/min^34,38^. In addition to the ambiguity surrounding the definition of high flow, many studies in the current literature did not quantify fistula flows but instead compared AVFs to central venous catheters or classified proximal AVFs as having higher flows than distal AVFs ^39-41^. Moreover, some studies excluded participants with AVF flow rates exceeding 2 L/min ^31^, further complicating interpretation of the findings. The relatively small number of participants with high-flow AVFs across studies may also have limited the ability to fully assess the impact of AVF flow on mortality. Similarly, the diagnosis of HF in dialysis patients is complex and overlaps with characteristic clinical features of patients with ESKD, such as dyspnoea on exertion. Moreover, the diagnosis of heart failure in ESKD lacks standardization. Adoption of heart failure diagnostic criteria as detailed in the 2014 ADQI proposal for a functional classification of heart failure in patients with ESKD would allow more accurate reporting of incident heart failure in this population ^42^. Finally, most of the included body of literature is at a high risk of bias. The multi-morbidity seen in patients with ESKD likely contributes to the development of HF and is difficult to control for in observational studies. This results in a significant risk of bias across most of the existing literature. Much of the current literature fails to identify confounders, such as pre-existing heart failure, and does not report which variables were controlled for making it difficult to establish the significance of findings.

This SWiM analysis suggests that there is a relationship between higher AVF flow and the incidence and severity of HF. While confirming the association between AVF flow and cardiovascular outcomes, this study highlights several key limitations that underscore the necessity for well-designed prospective investigations. Future studies should include clearly defined criteria for high-flow AVFs, as well as standardized diagnostic and staging protocols for HF in dialysis patients. Such studies should incorporate a comprehensive range of clinical, biochemical, and imaging data, with uniform timepoints for data collection and outcome assessment. We believe that without a concerted effort to standardize definitions and reporting outcomes in the dialysis population, a deeper understanding of the intricate relationship between AVF flow and cardiovascular outcomes will remain unattainable.

## Supporting information

PRISMA Checklist

## Data Availability Statement

All data used in this systematic review are publicly available from previously published studies. Citations and references for the included studies are provided within the manuscript and supplementary materials. Template data collection forms, data extracted from included studies, and data used for all analyses will be made available by the corresponding author upon request.

## References

1. Vanholder R, Annemans L, Brown E, et al. Reducing the costs of chronic kidney disease while delivering quality health care: a call to action. Nature Reviews Nephrology. 2017/07/01 2017;13(7):393–409. doi:10.1038/nrneph.2017.63

2. Tonelli M, Wiebe N, Culleton B, et al. Chronic kidney disease and mortality risk: a systematic review. J Am Soc Nephrol. Jul 2006;17(7):2034–47. doi:10.1681/ASN.2005101085

3. Webster AC, Nagler EV, Morton RL, Masson P. Chronic Kidney Disease. Lancet. Mar 25 2017;389(10075):1238–1252. doi:10.1016/S0140-6736(16)32064-5

4. Weiner DE, Tighiouart H, Elsayed EF, et al. The Framingham predictive instrument in chronic kidney disease. J Am Coll Cardiol. Jul 17 2007;50(3):217–24. doi:10.1016/j.jacc.2007.03.037

5. Mann JF, Gerstein HC, Pogue J, Bosch J, Yusuf S. Renal insufficiency as a predictor of cardiovascular outcomes and the impact of ramipril: the HOPE randomized trial. Ann Intern Med. Apr 17 2001;134(8):629–36. doi:10.7326/0003-4819-134-8-200104170-00007

6. Muntner P, He J, Hamm L, Loria C, Whelton PK. Renal insufficiency and subsequent death resulting from cardiovascular disease in the United States. J Am Soc Nephrol. Mar 2002;13(3):745–753. doi:10.1681/ASN.V133745

7. Go AS, Chertow GM, Fan D, McCulloch CE, Hsu CY. Chronic kidney disease and the risks of death, cardiovascular events, and hospitalization. N Engl J Med. Sep 23 2004;351(13):1296–305. doi:10.1056/NEJMoa041031

8. Hillege HL, Fidler V, Diercks GF, et al. Urinary albumin excretion predicts cardiovascular and noncardiovascular mortality in general population. Circulation. Oct 1 2002;106(14):1777–82. doi:10.1161/01.cir.0000031732.78052.81

9. Drueke TB, Massy ZA. Atherosclerosis in CKD: differences from the general population. Nat Rev Nephrol. Dec 2010;6(12):723–35. doi:10.1038/nrneph.2010.143

10. System USRD. 2021 USRDS Annual Data Report: Epidemiology of kidney disease in the United States., 2021. https://adr.usrds.org/2021

11. Registry. A. 44th Report, Chapter 3: Mortality in Kidney Failure with Replacement Therapy. 2021. Australia and New Zealand Dialysis and Transplant Registry. http://www.anzdata.org.au

12. Lok CE. Fistula first initiative: advantages and pitfalls. Clin J Am Soc Nephrol. Sep 2007;2(5):1043–53. doi:10.2215/CJN.01080307

13. Nakano J, Deschryver C. Effects of Arteriovenous Fistula on Systemic and Pulmonary Circulations. Am J Physiol. Dec 1964;207:1319–24. doi:10.1152/ajplegacy.1964.207.6.1319

14. Savage MT, Ferro CJ, Sassano A, Tomson CR. The impact of arteriovenous fistula formation on central hemodynamic pressures in chronic renal failure patients: a prospective study. Am J Kidney Dis. Oct 2002;40(4):753–9. doi:10.1053/ajkd.2002.35686

15. Reddy YNV, Obokata M, Dean PG, Melenovsky V, Nath KA, Borlaug BA. Long-term cardiovascular changes following creation of arteriovenous fistula in patients with end stage renal disease. Eur Heart J. Jun 21 2017;38(24):1913–1923. doi:10.1093/eurheartj/ehx045

16. Agarwal AK. Systemic Effects of Hemodialysis Access. Adv Chronic Kidney Dis. Nov 2015;22(6):459–65. doi:10.1053/j.ackd.2015.07.003

17. Stoumpos S, Rankin A, Hall Barrientos P, et al. Interrogating the haemodynamic effects of haemodialysis arteriovenous fistula on cardiac structure and function. Sci Rep. Sep 13 2021;11(1):18102. doi:10.1038/s41598-021-97625-5

18. Eriguchi M, Tsuruya K, Lopes M, et al. Routinely measured cardiac troponin I and N-terminal pro-B-type natriuretic peptide as predictors of mortality in haemodialysis patients. ESC Heart Fail. Apr 2022;9(2):1138–1151. doi:10.1002/ehf2.13784

19. Hu Z, Zhu F, Zhang N, et al. Impact of arteriovenous fistula blood flow on serum il-6, cardiovascular events and death: An ambispective cohort analysis of 64 Chinese hemodialysis patients. PLoS One. 2017;12(3):e0172490. doi:10.1371/journal.pone.0172490

20. Meyer-Olesen CL, Lindhard K, Jorgensen NR, et al. Flow reduction of a high-flow arteriovenous fistula in a hemodialysis patient reveals changes in natriuretic and renin-angiotensin system hormones of relevance for kidney function. Physiol Rep. Oct 2021;9(19):e14989. doi:10.14814/phy2.14989

21. Valerianova A, Malik J, Janeckova J, et al. Reduction of arteriovenous access blood flow leads to biventricular unloading in haemodialysis patients. Int J Cardiol. Jul 1 2021;334:148–153. doi:10.1016/j.ijcard.2021.04.027

22. Oe K, Araki T, Katano K, et al. Impact of inflow reduction of arteriovenous fistula on systemic hemodynamics in a patient with high-output heart failure during hemodialysis: a case report. Journal of Cardiology Cases. 2010;1(2):e98–e101.

23. Movilli E, Viola BF, Brunori G, et al. Long-term Effects of Arteriovenous Fistula Closure on Echocardiographic Functional and Structural Findings in Hemodialysis Patients: A Prospective Study. American Journal of Kidney Diseases. 2010/04/01/ 2010;55(4):682-689. doi:10.1053/j.ajkd.2009.11.008

24. Moher D, Liberati A, Tetzlaff J, Altman DG, Group P. Preferred reporting items for systematic reviews and meta-analyses: the PRISMA statement. PLoS Med. Jul 21 2009;6(7):e1000097. doi:10.1371/journal.pmed.1000097

25. Sterne JAC, Savović J, Page MJ, et al. RoB 2: a revised tool for assessing risk of bias in randomised trials. BMJ. 2019;366: l4898. doi:10.1136/bmj.l4898

26. Wells G, Shea B, O’connell D, et al. Newcastle-Ottawa quality assessment scale cohort studies. University of Ottawa. 2014;

27. Campbell M, McKenzie JE, Sowden A, et al. Synthesis without meta-analysis (SWiM) in systematic reviews: reporting guideline. BMJ. Jan 16 2020;368: l6890. doi:10.1136/bmj.l6890

28. Basile C, Vernaglione L, Casucci F, et al. The impact of haemodialysis arteriovenous fistulaon haemodynamic modifications of the cardiovascular system. Conference Abstract. Nephrology Dialysis Transplantation. May 2016;1):i259-i260. doi:10.1093/ndt/gfw173.2

29. Dundon BK, Torpey DK, Nelson AJ, et al. Beneficial cardiovascular remodeling following arterio-venous fistula ligation post-renal transplantation: a longitudinal magnetic resonance imaging study. Clin Transplant. Aug 2014;28(8):916–25. doi:10.1111/ctr.12402

30. Rao NN, Stokes MB, Rajwani A, et al. Effects of Arteriovenous Fistula Ligation on Cardiac Structure and Function in Kidney Transplant Recipients. Circulation. Jun 18 2019;139(25):2809–2818. doi:10.1161/CIRCULATIONAHA.118.038505

31. Wu CK, Wu CL, Lin CH, Leu JG, Kor CT, Tarng DC. Association of vascular access flow with short-term and long-term mortality in chronic haemodialysis patients: a retrospective cohort study. BMJ Open. Sep 24 2017;7(9):e017035. doi:10.1136/bmjopen-2017-017035

32. Yap YS, Chi WC, Lin CH, Liu YC, Wu YW. Association of early failure of arteriovenous fistula with mortality in hemodialysis patients. Sci Rep. Mar 11 2021;11(1):5699. doi:10.1038/s41598-021-85267-6

33. Hu Z, Wang P, Zeng R, Xu G. Impact of arteriovenous fistula blood flow on serum IL-6, cardiovascular events and death in Chinese patients receiving hemodialysis: A 5-year follow-up. Conference Abstract. Hong Kong Journal of Nephrology. October 2015;1):S135. doi:10.1016/j.hkjn.2015.09.236

34. Lok CE, Huber TS, Lee T, et al. KDOQI Clinical Practice Guideline for Vascular Access: 2019 Update. Am J Kidney Dis. Apr 2020;75(4 Suppl 2):S1–S164. doi:10.1053/j.ajkd.2019.12.001

35. Hetz P, Pirklbauer M, Muller S, Posch L, Gummerer M, Tiefenthaler M. Prophylactic Ligature of AV Fistula Prevents High Output Heart Failure after Kidney Transplantation. Am J Nephrol. 2020;51(7):511–519. doi:10.1159/000508957

36. Brouwers FP, de Boer RA, van der Harst P, et al. Incidence and epidemiology of new onset heart failure with preserved vs. reduced ejection fraction in a community-based cohort: 11-year follow-up of PREVEND. Eur Heart J. May 2013;34(19):1424–31. doi:10.1093/eurheartj/eht066

37. Borlaug BA, Kass DA. Ventricular-vascular interaction in heart failure. Cardiol Clin. Aug 2011;29(3):447–59. doi:10.1016/j.ccl.2011.06.004

38. Basile C, Lomonte C, Vernaglione L, Casucci F, Antonelli M, Losurdo N. The relationship between the flow of arteriovenous fistula and cardiac output in haemodialysis patients. Nephrol Dial Transplant. Jan 2008;23(1):282–7. doi:10.1093/ndt/gfm549

39. Chuang MK, Chang CH, Chan CY. The effect of haemodialysis access types on cardiac performance and morbidities in patients with symptomatic heart disease. PLoS ONE. 01 Feb 2016;11(2) (no pagination)e0148278. doi:10.1371/journal.pone.0148278

40. Drouven JW, Wiegersma J, Assa S, et al. Differences in shuntflow (Qa), cardiac function and mortality between hemodialysis patients with a lower-arm fistula, an upper-arm fistula, and an arteriovenous graft. J Vasc Access. Apr 23 2022:11297298221092741. doi:10.1177/11297298221092741

41. Malyszko J, Kozminski P, Malyszko J, Mysliwiec M. Type of arteriovenous fistula, NYHA class and apelin in hemodialyzed patients. Int Urol Nephrol. Mar 2011;43(1):185–90. doi:10.1007/s11255-009-9667-1

42. Chawla LS, Herzog CA, Costanzo MR, et al. Proposal for a functional classification system of heart failure in patients with end-stage renal disease: proceedings of the acute dialysis quality initiative (ADQI) XI workgroup. J Am Coll Cardiol. Apr 8 2014;63(13):1246–1252. doi:10.1016/j.jacc.2014.01.020

