## Supplemental Tables for "A Change of Heart: The Effect of High Flow Arteriovenous Fistulas on Cardiovascular Outcomes - a Systematic Review and Synthesis without Meta-analysis (SWiM)"

### Supplementary Tables

| Supplementary Table 1: Database Search Strategy | |
|---|---|
| **Database** | **Search Strategy** |
| Pubmed | ((dialysis access* [tiab]) OR (vascular access* [tiab]) OR (hemodialysis access* [tiab]) OR (haemodialysis access* [tiab]) OR (arteriovenous fistula* [tiab]) OR (arteriovenous shunt* [tiab]) OR (AVF* [tiab]) OR (surgical shunt* [tiab])) AND ((cardiac structure [tiab]) OR (cardiac function [tiab]) OR (heart failure [tiab]) OR (cardiac failure [tiab]) OR (diastolic failure [tiab]) OR (systolic failure [tiab]) OR (diastolic dysfunction [tiab]) OR (systolic dysfunction [tiab]) OR (HFpEF [tiab]) OR (HFrEF [tiab])). |
| Cochrane Central Register of Controlled Trials (CENTRAL) | ((dialysis access*) OR (vascular access*) OR (hemodialysis access*) OR (haemodialysis access*) OR (arteriovenous fistula*) OR (arteriovenous shunt*) OR (AVF*) OR (surgical shunt*)) AND (HF?EF OR ((Cardiac or heart) adj (failure or dysfunction or structure or function or index or output)) OR ((systolic or diastolic) adj (failure or dysfunction))). |
| Embase(Ovid) | (HF?EF.tw.OR ((Cardiac or heart) adj (failure or dysfunction or structure or function or index or output)).tw. OR ((systolic or diastolic) adj (failure or dysfunction)).tw.) AND ((dialysis or vascular or arteriovenous or AV? Or surgical) adj (access* or fistula* or shunt*)).tw. |

| **Supplementary Table 2: Summary of Outcome Groupings for SWiM Analysis.** Comparator Group: High vs. Low AVF flow as defined by the study, Standardization of metrics for each outcome: direction of effect, Alternative synthesis method employed: vote counting of direction of effect, Study prioritization criteria: No study prioritization. | | | | | | |
|---|---|---|---|---|---|---|
| **Outcome Category: Group** | **Details of Study Grouping** | **Author** | **Year** | **Journal** | **PMID** | **Reference** |
| Clinical: Incident HF | Incident HF | Hetz, P | 2020 | Am J Nephrology | 632409326 | 1 |
| | | Kaller, R | 2023 | J Clin Med | 37445452 | 2 |
| | | Martinez-Gallardo, R | 2012 | Nefrologia | 365455836 | 3 |
| | | Reddy, YNV | 2017 | Eur Heart Journal | 617006398 | 4 |
| | | Shah, NA | 2024 | J Vasc Surg | 39631475 | 5 |
| | | Stoumpos, S | 2024 | Clin Kidney J | 38737344 | 6 |
| Clinical: HF Composite Outcome | Prevalent HF, change in HF symptom burden | Abbott, KC | 2003 | J Nephrology | 14736009 | 7 |
| | | Basile, C | 2008 | Neph Dial Trans | 17942475 | 8 |
| | | Blanchard, V | 2020 | Heart & Vessels | 200510969 | 9 |
| | | Chuang, MK | 2016 | PLoS ONE | 608547220 | 10 |
| | | Keller, N | 2021 | Sem in Dialysis | — | 11 |
| | | Lee, DY | 2024 | ESC Heart Fail | 37885349 | 12 |
| | | Letachowicz, K | 2021 | Renal Failure | 33397180 | 13 |
| | | Malin, J | 2024 | Heart Lung | 39116576 | 14 |
| | | Malyszko, J | 2011 | Int Urol Nephrol | 19908160 | 15 |
| | | Malyszko, J | 2011 | Renal Failure | 362800652 | 16 |
| | | Reddy, YNV | 2017 | Euro Heart J | 617006398 | 4 |
| | | Yu, Q | 2011 | J Nephrology | 362536606 | 17 |
| | | Zamboli, P | 2018 | J Nephrology | 29357085 | 18 |
| Clinical: improvement in HF symptom burden with Flow Reduction | Flow reduction intervention (AVF banding, AVF ligation, etc) | Alqassieh, A | 2021 | American Surg | 34794331 | 19 |
| | | Balamuthsamy, S | 2016 | HD International | 26663664 | 20 |
| | | Gkotsis, G | 2016 | Annals Vasc Surg | — | 21 |
| | | Haque, A | 2022 | Annals Vasc Surg | 2015987935 | 22 |
| | | Huang, XM | 2023 | J Vasc Surg | 37086822 | 23 |
| | | Janeckova, J | 2024 | Transpl Int | 39188270 | 24 |
| | | Kanno, T | 2015 | J Vasc Access | — | 25 |
| | | Kurita, N | 2011 | Ther Apheresis Dial | — | 26 |
| | | Lee, H | 2020 | Vasc Endovasc Surg | 31506033 | 27 |
| | | Miller, GA | 2010 | Int Soc Nephrol | — | 28 |
| | | Schier, T | 2013 | Clin Transplant | 52817589 | 29 |
| | | Zanow, J | 2006 | J Vasc Surg | 17145429 | 30 |

| | | | | | | |
|---|---|---|---|---|---|---|
| Clinical: All-cause mortality | All-cause mortality | Drouven, JW | 2022 | J Vasc Access | 35466801 | 31 |
| | | Gan, W | 2023 | J Vasc Access | 37997027 | 32 |
| | | Hu, Z | 2017 | PLoS ONE | — | 33 |
| | | Quiroga, B | 2013 | Blood Purification | 603979658 | 34 |
| | | Reddy, YNV | 2017 | Euro Heart J | 617006398 | 4 |
| | | Repasos, E | 2017 | Hellenic J Cardiol | 619763439 | 35 |
| | | Stoumpos, S | 2024 | Clin Kidney J | 38737344 | 6 |
| | | Wu, CK | 2017 | BMJ Open | 618455534 | 36 |
| | | Yu, Y | 2020 | Blood Purification | 62958285 | 37 |

| | | | | | | |
|---|---|---|---|---|---|---|
| Other: General Cardiac Performance Parameters | CO, CI | Aitken, E | 2020 | Heart and Vessels | 25634156 | 38 |
| | | Basile, C | 2016 | Clin Kidney J | 27679720 | 39 |
| | | Blanchard, V | 2015 | J Vasc Access | 2005109691 | 9 |
| | | Bos, WJW | 1999 | Kidney International | 29197995 | 40 |
| | | Chemla, ES | 2007 | Seminars in Dialysis | 46144007 | 41 |
| | | Choi, YS | 2022 | Front Cardiovasc Med | 35966517 | 42 |
| | | Cridlig, J | 2008 | Transplant Int | 352316411 | 43 |
| | | Dongradi, G | 1981 | Clinical Nephrology | 7214756 | 44 |
| | | Dundon, BK | 2014 | Clin Transplant | 24931318 | 45 |
| | | Dundon, BK | 2014 | Int J Nephrol and RV Disease | 25258554 | 46 |
| | | Elkin, DC | 1947 | JAMA | 20255627 | 47 |
| | | Hashimoto, K | 2021 | Science Progress | 34281440 | 48 |
| | | Huang, XM | 2023 | J Vasc Surg | 37086822 | 23 |
| | | Iwashima, Y | 2002 | Am J of Kidney Dis | 35265477 | 49 |
| | | Janeckova, J | 2024 | Transpl Int | 39188270 | 24 |
| | | Keller, N | 2021 | Seminars in Dialysis | — | 11 |
| | | Malik, J | 2021 | ESC Heart Failure | 2010875987 | 50 |
| | | Malin, J | 2024 | Heart Lung | 39116576 | 14 |
| | | Meier, M | 2010 | Transplant Int | 358395077 | 51 |
| | | Ogugua, FM | 2024 | Echocardiography | 38113338 | 52 |
| | | Ori, Y | 1996 | NDT | 26038471 | 53 |
| | | Payne RM | 1972 | Kidney Int | — | 54 |
| | | Polkinghorne, KR | 2004 | Nephrology | 39318643 | 55 |
| | | Prastowo, RA | 2022 | Annals of Med & Surg | 2017822623 | 56 |
| | | Rao, NN | 2019 | Circulation | 629641077 | 57 |
| | | Reddy, YNV | 2017 | Euro Heart Journal | 617006398 | 4 |
| | | Samet, P | 1964 | Am J Cardiology | 281112168 | 58 |
| | | Sandhu, JS | 2004 | Renal Failure | 39579132 | 59 |
| | | Saratzis, AN | 2008 | J of Vasc Access | 354667983 | 60 |
| | | Saritas, B | 2010 | Turkiye Klinikleri CV Sci | 359540502 | 61 |
| | | Sheashaa, H | 2004 | American J of Nephrol | 39279386 | 62 |
| | | Tang, CY | 2019 | Therapeutic Apher Dial | 628158694 | 63 |
| | | Tellioglu, G | 2008 | Transplantation Proc | 351186655 | 64 |
| | | Unger, P | 2008 | Transplantation | 351435955 | 65 |
| | | Valek, M | 2010 | ASAIO journal | 359330420 | 66 |
| | | Velez-Roa, S | 2004 | Nephro Dial Transp | 38786758 | 67 |
| | | Warren, JV | 1947 | JCI | 280583947 | 68 |
| | | Warren, JV | 1951 | JCI | 280936836 | 69 |
| | | Wijnen, E | 2005 | Artificial Organs | 43943031 | 70 |
| | | Ye, WL | 2013 | Acta Acad Med Sinicae | 368691545 | 71 |
| | | Zamboli , P | 2018 | J Nephrol | 29357085 | 18 |

| | | | | | | |
|---|---|---|---|---|---|---|
| | | Glowinski, J | 2012 | Pols Arch Med Wewen | — | 72 |
| | | Hiremath, S | 2010 | Nephro Dial Trans | 359210004 | 73 |
| | | He, X | 2023 | Pak J Med Sci | 37250573 | 74 |
| | | Huang, XM | 2023 | J Vasc Surg | 37086822 | 23 |
| | | Jaques, DA | 2021 | Nephro Dial Trans | — | 75 |
| | | Keller, N | 2021 | Sem in Dialysis | — | 11 |
| | | Kurita, N | 2011 | Ther Apheres & Dial | — | 26 |
| | | Laranjinha, I | 2018 | J Brasil de Nefro | — | 76 |
| | | Movilli, E | 2010 | Am J of Kidney Dis | — | 77 |
| TTE: Structural Parameters | LVM, LVMI | Reddy, YNV | 2017 | Euro Heart J | 617006398 | 4 |
| | | Saleh, MA | 2018 | Egypt Heart J | 200128804 | 78 |
| | | Salehi, T | 2021 | Kidney 360 | CN02390203 | 79 |
| | | Sheashaa, H | 2004 | Am J of Nephrol | 39279386 | 62 |
| | | Song, W | 2024 | Front Cardiovasc Med | 38682103 | 80 |
| | | Unger, P | 2002 | Transplantation | 12134102 | 81 |
| | | Unger, P | 2008 | Transplantation | 351435955 | 65 |
| | | Valerianova, A | 2021 | Int J of Cardiology | 2011890042 | 82 |
| | | Van Duijnhoven, ECM | 2001 | Neph Dial Trans | 32123928 | 83 |
| | | Wohlfahrt, P | 2016 | HTN Research | 612013784 | 84 |

| | | | | | | |
|---|---|---|---|---|---|---|
| TTE: LV Systolic Function | EF | Balamuthusamy, S | 2016 | Hemodialysis Int | 26663664 | 20 |
| | | Blanchard, V | 2020 | Heart & Vessels | 2005109691 | 9 |
| | | Beigi, AA | 2009 | J Vasc Access | 355609373 | 85 |
| | | Chen, R | 2014 | PLoS ONE | 373418966 | 86 |
| | | Chuang, MK | 2016 | PLoS ONE | 608547220 | 10 |
| | | Cridlig, J | 2008 | Transp Int | 352316411 | 43 |
| | | De Lima, JJG | 1999 | Cardiology | CN-00297255 | 87 |
| | | Di Lullo, L | 2011 | Nephron Clin Practice | 51214731 | 88 |
| | | Glowinski, J | 2012 | Pol Arch Med Wewne | 365615736 | 72 |
| | | Hashimoto, K | 2021 | Science Prog | 34281440 | 48 |
| | | He, X | 2023 | Pak J Med Sci | 37250573 | 74 |
| | | Huang, XM | 2023 | J Vasc Surg | 37086822 | 23 |
| | | Jaques, DA | 2021 | Nephrol Dial Transp | — | 75 |
| | | Keller, N | 2021 | Sem in Dialysis | — | 11 |
| | | Kurita, N | 2011 | Ther Apheres & Dial | — | 26 |
| | | Laranjinha, I | 2018 | J Bras de Nefrologia | — | 76 |
| | | Malik, J | 2021 | ESC Heart Fail | 2010875987 | 50 |
| | | Malin, J | 2024 | Heart Lung | 39116576 | 14 |
| | | Malyszko, J | 2011 | Int Urol Nephrol | 19908160 | 16 |
| | | Malyszko, J | 2011 | Renal Failure | 362800652 | 15 |
| | | Meier, M | 2010 | Transplant Int | 358395077 | 51 |
| | | Movilli, E | 2010 | Am J of Kidney Disease | — | 77 |
| | | Ogugua, FM | 2024 | Echocardiograph | 38113338 | 52 |
| | | Ori, Y | 1996 | Nephrol Dial Transp | 26038471 | 53 |
| | | Paneni, F | 2013 | Journal of CV Medicine | — | 89 |
| | | Papasotiriou, M | 2019 | Exp & Clin Transp | 2002736460 | 90 |
| | | Prastowo, RA | 2022 | Annals of Med & Surg | 2017822623 | 56 |
| | | Reddy, YNV | 2017 | Euro Heart J | 617006398 | 4 |
| | | Said, K | 2018 | Heart Lung Circ | 29866523 | 91 |
| | | Saleh, MA | 2018 | Egypt Heart J | 200128804 | 78 |
| | | Salehi, T | 2021 | Kidney 360 | CN02390203 | 79 |
| | | Saritas, B | 2010 | Truk Klin CV Sci | 359540502 | 61 |
| | | Sheashaa, H | 2004 | Am J of Nephrol | 39279386 | 62 |
| | | Song, W | 2024 | Front Cardiovasc Med | 38682103 | 80 |
| | | Tayebi, P | 2024 | Vasc Specialist Int | 38454861 | 92 |
| | | Unger, P | 2008 | Transplantation | 351435955 | 65 |
| | | Unger, P | 2002 | Transplantation | 12134102 | 81 |
| | | Ye, WL | 2013 | Acta Acad Med Sinic | 3686915545 | 71 |
| | | Zamboli, P | 2018 | J Nephrol | 29357085 | 18 |

| | | | | | | |
|---|---|---|---|---|---|---|
| TTE: Diastolic Function* | E/e', LAVI, TR Velocity, Septal or Lateral e’ Velocity, E/A Ratio<br><br>**each included study had to report at least 2 parameters of diastolic function with all reported parameters noting the same direction of effect to be counted as either a positive or negative study. If reported parameters had differing directions of effect, the study was counted as a neutral study.* | Abdelwhab, S | 2008 | Am J of Nephrol | 18635926 | 93 |
| | | Blanchard, V | 2020 | Heart and Vessels | 2005109691 | 9 |
| | | Chen, R | 2014 | PLoS ONE | 373418966 | 86 |
| | | Cridlig, J | 2008 | Transp Int | 352316411 | 43 |
| | | Di Lullo, L | 2011 | Nephron Clin Practice | 51214731 | 88 |
| | | Drouven, JW | 2023 | J Vasc Access | 35466801 | 31 |
| | | Huang, XM | 2023 | J Vasc Surg | 37086822 | 23 |
| | | Iwashima, Y | 2002 | Am J of Kidney Dis | 35265477 | 49 |
| | | Jaques, DA | 2021 | Nephrol Dial Transp | — | 75 |
| | | Keller, N | 2022 | Seminars in Dial | — | 11 |
| | | Ori, Y | 2002 | Am J of Kidney Dis | 35101329 | 94 |
| | | Paneni, F | 2013 | J of CVS Med | — | 89 |
| | | Prastowo, RA | 2022 | Annals Med Surg | 2017822623 | 56 |
| | | Reddy, YNV | 2017 | Eur Heart J | 617006398 | 4 |
| | | Said, K | 2018 | Heart Lung Circ | 29866523 | 91 |
| | | Salehi, T | 2021 | Kidney 360 | CN-02390203 | 79 |
| | | Ye, WL | 2013 | Acta Acad Med Sinic | 3686915545 | 71 |
| TTE: Pulmonary Hypertension | Pulmonary HTN, PAP, PASP | Abdelghany, MF | 2020 | Egyp J Chest and Tb | — | 95 |
| | | Abdelwhab, S | 2008 | Am J Nephrol | 18635926 | 93 |
| | | Acarturk, G | 2008 | Int Urol and Neph | 17985208 | 96 |
| | | Ayyaz, A | 2020 | Paki J of Med Health Sci | 2010623597 | 97 |
| | | Balamuthusamy, S | 2016 | Haemodialysis Int | 26663664 | 20 |
| | | Beigi, AA | 2009 | J Vasc Access | 355609373 | 85 |
| | | Cridlig, J | 2008 | Transp Int | 352316411 | 43 |
| | | Di Lullo, L | 2011 | Nephron Clin Practice | 51214731 | 88 |
| | | Emara, MM | 2013 | Egyp J Chest and Tb | 370426387 | 98 |
| | | Havlucu, Y | 2007 | Respiration | — | 99 |
| | | Huang, XM | 2023 | J Vasc Surg | 37086822 | 23 |
| | | Keller, N | 2022 | Sem in Dial | — | 11 |
| | | Laranjinha, I | 2018 | J Brasil de Nefro | — | 76 |
| | | Malik, J | 2021 | ESC Heart Failure | 2010875987 | 50 |
| | | Meier, M | 2010 | Transp Int | 358395077 | 51 |
| | | Ogugua, FM | 2024 | Echocardiography | 38113338 | 52 |
| | | Reque, J | 2017 | Am J Nephrology | — | 100 |
| | | Said, K | 2018 | Heart Lung Circ | 29866523 | 91 |
| | | Saleh, MA | 2018 | Egypt Heart J | 2001288044 | 78 |
| | | Song, W | 2024 | Front Cardiovasc Med | 38682103 | 80 |
| | | Tudoran, M | 2020 | Niger J Clin Pract | 32031094 | 101 |
| | | Valerianova, A | 2021 | Int J of Card | 2011890042 | 82 |

| | | | | | | |
|---|---|---|---|---|---|---|
| | | Warner, ED | 2024 | Curr Probl Cardiol | 38237814 | 102 |
| | | Wohlfahrt, P | 2016 | HTN Research | 612013784 | 84 |
| | | Ye, WL | 2013 | Act Acad Med Sini | 368691545 | 71 |
| | | Yigla, M | 2003 | Chest | 36549778 | 103 |
| | | Zamboli, P | 2018 | J Nephrol | 29357085 | 18 |
| | | Zhao, L | 2014 | Sich da xue xue bao | 25341347 | 104 |
| Biomarkers: Inflammatory | CRP, IL-6 | Hu, Z | 2017 | PLoS ONE | — | 33 |
| | | Kaller, R | 2023 | J Clin Med | 37445452 | 2 |
| | | Malyszko, J | 2011 | Int Urol Nephrol | 19908160 | 16 |
| | | Malyszko, J | 2011 | Renal Failure | 362800652 | 15 |
| | | Stoumpos S | 2024 | Clin Kidney J | 38737344 | 6 |
| Biomarkers: Volume Load | BNP, NT-ProBNP, ANP | Abdelwhab, S | 2008 | AM J Nephrol | 18635926 | 93 |
| | | Adachi, T | 2016 | Hemodialysis Int | 27669543 | 105 |
| | | Blanchard, V | 2020 | Heart and Vessels | 2005109691 | 9 |
| | | Choi, YS | 2022 | Front Cardiovasc Med | 35966517 | 42 |
| | | Haque, A | 2022 | Annals of Vasc Surg | 2015987935 | 22 |
| | | Hiremath, S | 2010 | NDT | 359210004 | 73 |
| | | Iwashima, Y | 2002 | AJKD | — | 49 |
| | | Jaques, DA | 2021 | NDT | — | 75 |
| | | Keller, N | 2022 | Seminars in Dialysis | — | 11 |
| | | Malik, J | 2009 | Int Urol Nephrol | 362800652 | 106 |
| | | Malyszko, J | 2011 | Int Urol Nephrol | 19908160 | 16 |
| | | Ori, Y | 1996 | NDT | 26038471 | 53 |
| | | Rao, NN | 2019 | Circulation | 629641077 | 57 |
| | | Schier, T | 2013 | Clinical Transplantation | 52817589 | 29 |
| | | Stoumpos, S | 2021 | Scientific Reports | 636137643 | 107 |

**Supplementary Table 3: Newcastle-Ottawa Risk of Bias Assessment of Observational Studies**

| Author | Year | Journal | Participants (n) | Selection | | | | Comparability | | Outcome | | | Risk of Bias |
|---|---|---|---|---|---|---|---|---|---|---|---|---|---|
| | | | | Representativeness of Exposure cohort | Non-exposure | Ascertainment of Exposure | Outcome of Interest Absent at start of study | Primary study controls | Secondary Study Controls | Assessment of Outcome | Follow-up: Time | Follow Up: Cohort | |
| Abbott, KC | 2003 | J Nephrol | 993 | a | b | a | b | c | c | b | a | a | Poor |
| Aitken, E | 2015 | J Vasc Access | 100 | a | a | a | b | c | c | b | a | a | Poor |
| Alqassieh, A | 2021 | Am Surg | 7 | b | a | a | a | c | c | c | a | b | Poor |
| Balamuthusamy, S | 2016 | Hemodialysis International | 12 | b | a | a | b | c | c | b | a | a | Poor |
| Basile, C | 2008 | Nephrol Dial Transplant | 96 | a | a | a | b | c | c | b | a | a | Poor |
| Basile, C | 2016 | Clinical Kidney J | 86 | a | a | a | b | c | c | d | a | d | Poor |
| Blanchard, V | 2020 | Heart and Vessels | 47 | a | a | a | b | b | b | b | a | a | Poor |
| Chemla, ES | 2007 | Seminars in Dialysis | 17 | a | a | a | b | b | b | b | a | a | Poor |
| Choi, YS | 2022 | Front Cardiovasc Med | 35 | a | b | a | a | c | c | d | a | a | Fair |
| Chuang, M | 2016 | PLoS ONE | 103 | b | a | a | a | a | a | b | a | a | Poor |
| Dongradi, G | 1981 | Clinical Nephrology | 16 | a | a | a | a | a | a | b | a | a | Good |
| Drouven, J | 2023 | J Vasc Access | 100 | a | a | a | a | b | b | b | a | a | Good |
| Gan, W | 2023 | J Vasc Access | 261 | a | a | a | a | c | c | b | a | b | Good |
| Gkotsis, G | 2016 | Annals of Vasc Surg | 12 | a | a | a | a | a | a | b | a | a | Good |
| Haque, A | 2022 | Annals of Vasc Surg | 11 | a | a | a | a | a | a | b | a | a | Good |
| He, X | 2023 | Pak J Med Sci | 270 | a | a | a | a | c | c | b | a | d | Good |
| Hu, Z | 2017 | PLoS ONE | 64 | a | a | a | b | c | c | b | a | a | Poor |
| Huang, XM | 2023 | J Vasc Surg | 31 | a | a | a | a | c | c | a | a | a | Good |
| Janeckova, J | 2024 | Transpl Int | 40 | a | a | a | a | c | c | a | a | d | Good |
| Kaller, R | 2023 | J Clin Med | 42 | a | a | a | a | a | a | a | a | a | Good |
| Kanno, T | 2015 | J Vascur Access | 74 | b | a | a | b | c | c | b | b | a | Poor |
| Keller, N | 2021 | Seminars in Dialysis | 49 | b | a | a | b | c | c | b | a | a | Poor |
| Kurita, N | 2011 | Therapeutic Apheresis and Dial | 33 | b | a | a | a | c | c | b | a | d | Poor |

| | | | | | | | | | | | | | |
|---|---|---|---|---|---|---|---|---|---|---|---|---|---|
| Lee, D | 2024 | ESC Heart Fail | 109 | b | b | a | a | c | c | a | a | d | Fair |
| Lee, H | 2020 | Vascular and Endovascular Surgery | 5 | b | a | a | b | c | c | d | a | a | Poor |
| Letachowicz, K | 2021 | Renal Failure | 284 | a | a | a | b | b | b | c | a | b | Good |
| Malin, J | 2024 | Heart Lung | 651 | a | b | a | a | a | a | c | a | d | Good |
| Malyszko, J | 2011 | Int Urol Nephrol | 100 | a | a | a | b | c | c | d | a | a | Poor |
| Malyszko, J | 2011 | Renal Failure | 93 | a | a | a | b | c | c | d | a | a | Poor |
| Martinez-Gallardo, R | 2012 | Nefrologia | 562 | b | a | a | b | b | b | d | a | d | Poor |
| Miller, G | 2010 | Int Society of Nephrol | 69 | d | d | a | a | c | c | d | a | a | Poor |
| Ogugua, FM | 2024 | Echocardiography | 94 | a | a | a | a | a | a | b | a | d | Good |
| Quiroga, B | 2013 | Blood Purification | 211 | b | b | d | b | b | b | b | a | d | Fair |
| Quiroga, B | 2013 | Blood Purification | 211 | a | a | d | a | b | b | b | a | c | Good |
| Reddy, YNV | 2017 | European Heart Journal | 137 | a | a | a | a | b | b | b | a | a | Good |
| Repasos, E | 2017 | Hellenic Journal of Cardiology | 40 | b | a | a | a | b | b | b | a | a | Good |
| Salehi, T | 2021 | Kidney 360 | 45 | b | a | a | a | b | b | b | a | a | Good |
| Sandhu, JS | 2004 | Renal Failure | 8 | a | a | a | a | c | c | b | b | c | Poor |
| Saratzis , AN | 2008 | J Vasc Access | 10 | a | a | c | a | a | c | c | a | a | Good |
| Schier, T | 2013 | Clin Transplant | 113 | a | a | a | a | c | c | d | a | d | Fair |
| Shah, NA | 2024 | J Vasc Surg | 366 | a | a | a | a | a | a | a | a | d | Good |
| Song, W | 2024 | Front Cardiovasc Med | 153 | a | a | a | a | a | a | d | a | d | Good |
| Stoumpos, S | 2024 | Clin Kidney J | 1330 | a | a | a | a | a | a | a | a | a | Good |
| Tayebi, P | 2024 | Vasc Specialist Int | 100 | a | a | a | a | c | c | d | a | d | Fair |
| Tellioglu, G | 2008 | Transplantation Proceedings | 30 | a | a | a | b | a | a | d | a | d | Good |
| Tordoir, JHM | 1990 | Euro J of Vasc Surf | 90 | a | a | a | b | c | c | d | a | d | Poor |
| Salehi, M | 2018 | Egyptian Heart Journal | 100 | a | a | c | a | c | c | c | a | a | Poor |
| Warner, ED | 2024 | Curr Probl Cardiol | 478896 | a | a | a | b | a | a | b | a | a | Good |
| Wu, CK | 2017 | BMJ Open | 378 | a | a | a | a | b | b | b | a | b | Good |
| Yu, Y | 2020 | Blood Purification | 358 | b | a | a | a | b | b | a | a | c | Fair |
| Yu, Q | 2011 | Journal of Nephrology | 376 | c | a | a | b | b | b | b | a | d | Poor |
| Zamboli, P | 2018 | J Nephrol | 19 | b | a | a | b | c | c | b | a | a | Poor |
| Zanow, J | 2006 | J Vasc Surgery | 17 | d | d | a | b | c | c | d | a | a | Poor |
| Pandeya, S | 1999 | ASAIO journal | 16 | d | d | a | b | c | c | b | a | a | Poor |

Supplementary Table 4: Cochrane Risk-of-Bias Tool for Randomised Control Trials.

| Author | Year | Journal | Participants (n) | **Domain 1:** Randomization Process | **Domain 2:** Deviations from Interventions | **Domain 3:** Missing Outcome Data | **Domain 4:** Measurement of Outcome | **Domain 5:** Selection of Reported Result | Risk of Bias |
|---|---|---|---|---|---|---|---|---|---|
| Hetz, P | 2020 | American Journal of Nephrology | 28 | Some | Some | High | Low | Low | High |

References:


1. Hetz P, Pirklbauer M, Muller S, Posch L, Gummerer M, Tiefenthaler M. Prophylactic Ligature of AV Fistula Prevents High Output Heart Failure after Kidney Transplantation. *American Journal of Nephrology*. 01 Jul 2020;51(7):511-519. doi:https://dx.doi.org/10.1159/000508957
2. Kaller R, Russu E, Muresan AV, et al. Intimal CD31-Positive Relative Surfaces Are Associated with Systemic Inflammatory Markers and Maturation of Arteriovenous Fistula in Dialysis Patients. *Journal of Clinical Medicine*. 01 Jul 2023;12(13) (no pagination)4419. doi:https://dx.doi.org/10.3390/jcm12134419
3. Martinez-Gallardo R, Ferreira-Morong F, Garcia-Pino G, Cerezo-Arias I, Hernandez-Gallego R, Caravaca F. Congestive heart failure in patients with advanced chronic kidney disease: association with pre-emptive vascular access placement. *Nefrologia : publicacion oficial de la Sociedad Espanola Nefrologia*. 2012;32(2):206-212.
4. Reddy YNV, Obokata M, Dean PG, Melenovsky V, Nath KA, Borlaug BA. Long-term cardiovascular changes following creation of arteriovenous fistula in patients with end stage renal disease. *European Heart Journal*. 21 Jun 2017;38(24):1913-1923. doi:https://dx.doi.org/10.1093/eurheartj/ehx045
5. Shah NA, Byrne P, Endre ZH, Cochran BJ, Barber TJ, Erlich JH. Predicting high-flow arteriovenous fistulas and cardiac outcomes in hemodialysis patients. In Press. *Journal of Vascular Surgery*. 2025;doi:https://dx.doi.org/10.1016/j.jvs.2024.11.028
6. Stoumpos S, Van Rhijn P, Mangion K, Thomson PC, Mark PB. Arteriovenous fistula for haemodialysis as a predictor of de novo heart failure in kidney transplant recipients. *Clin Kidney J*. May 2024;17(5):sfae105. doi:10.1093/ckj/sfae105
7. Abbott KC, Trespalacios FC, Agodoa LY. Arteriovenous fistula use and heart disease in long-term elderly hemodialysis patients: analysis of United States Renal Data System Dialysis Morbidity and Mortality Wave II. *J Nephrol*. Nov-Dec 2003;16(6):822-30.
8. Basile C, Lomonte C, Vernaglione L, Casucci F, Antonelli M, Losurdo N. The relationship between the flow of arteriovenous fistula and cardiac output in haemodialysis patients. *Nephrol Dial Transplant*. Jan 2008;23(1):282-7. doi:10.1093/ndt/gfm549
9. Blanchard V, Courtellemont C, Cariou E, et al. Cardiac impact of arteriovenous fistulas: what tools to assess? *Heart and Vessels*. 01 Nov 2020;35(11):1583-1593. doi:https://dx.doi.org/10.1007/s00380-020-01630-z
10. Chuang MK, Chang CH, Chan CY. The effect of haemodialysis access types on cardiac performance and morbidities in patients with symptomatic heart disease. *PLoS ONE*. 01 Feb 2016;11(2) (no pagination)e0148278. doi:https://dx.doi.org/10.1371/journal.pone.0148278
11. Keller N, Monnier A, Caillard S, et al. High-flow arteriovenous fistula and hemodynamic consequences at 1year after kidney transplantation. *Seminars in dialysis*. 01 Mar 2022;35(2):171-180. doi:https://dx.doi.org/10.1111/sdi.13028
12. Lee DY, Chen T, Huang WC, et al. Systemic vascular resistance predicts high-output cardiac failure in patients with high-flow arteriovenous fistula. *ESC Heart Fail*. Feb 2024;11(1):189-197. doi:10.1002/ehf2.14563
13. Letachowicz K, Bardowska K, Krolicki T, et al. The impact of location and patency of the arteriovenous fistula on quality of life of kidney transplant recipients. *Renal Failure*. 2021;43(1):113-122. doi:https://dx.doi.org/10.1080/0886022X.2020.1865171
14. Malin J, Khan R, Manzano JMM, et al. Association of arteriovenous fistulae with precapillary pulmonary hypertension - A single center retrospective analysis of invasive hemodynamic parameters. *Heart Lung*. Nov-Dec 2024;68:260-264. doi:10.1016/j.hrtlng.2024.08.007
15. Malyszko J, Levin-Iaina N, Malyszko JS, Kozminski P, Koc-Zorawska E, Mysliwiec M. Copeptin and its relation to arteriovenous fistula (AVF) type and NYHA class in hemodialysis patients. *Renal Failure*. November 2011;33(10):929-934. doi:https://dx.doi.org/10.3109/0886022X.2011.618904

16. Malyszko J, Kozminski P, Malyszko J, Mysliwiec M. Type of arteriovenous fistula, NYHA class and apelin in hemodialyzed patients. *Int Urol Nephrol*. Mar 2011;43(1):185-90. doi:10.1007/s11255-009-9667-1
17. Yu Q, Yu H, Huang J, Chen S, Wang L, Yuan W. Distribution and complications of native arteriovenous fistulas in maintenance hemodialysis patients: A single-center study. *Journal of Nephrology*. September-Octember 2011;24(5):597-603. doi:https://dx.doi.org/10.5301/JN.2011.6251
18. Zamboli P, Borrelli S, Garofalo C, et al. High-flow arteriovenous fistula and heart failure: could the indexation of blood flow rate and echocardiography have a role in the identification of patients at higher risk? *Journal of Nephrology*. 01 Dec 2018;31(6):975-983. doi:https://dx.doi.org/10.1007/s40620-018-0472-8
19. Alqassieh A, Dennis PB, Mehta V, et al. MILLER Banding Procedure for Treatment of Dialysis Access-Related Steal Syndrome, Pulmonary Hypertension, and Heart Failure. *Am Surg*. Nov 18 2021:31348211056259. doi:10.1177/00031348211056259
20. Balamuthusamy S, Jalandhara N, Subramanian A, Mohanaselvan A. Flow reduction in high-flow arteriovenous fistulas improve cardiovascular parameters and decreases need for hospitalization. *Hemodial Int*. Jul 2016;20(3):362-8. doi:10.1111/hdi.12387
21. Gkotsis G, Jennings WC, Malik J, Mallios A, Taubman K. Treatment of High Flow Arteriovenous Fistulas after Successful Renal Transplant Using a Simple Precision Banding Technique. *Annals of Vascular Surgery*. 01 Feb 2016;31:85-90. doi:https://dx.doi.org/10.1016/j.avsg.2015.08.012
22. Haque A, Al-Khaffaf H. Early Results Using N-Terminal pro-B-Type Natriuretic Peptide (pro-BNP) as a Biomarker for the Efficacy of Secondary Extension Technique (SET) in Improving Myocardial Function in Dialysis Patients With High Flow Fistulas. *Annals of Vascular Surgery*. April 2022;81:267-272. doi:https://dx.doi.org/10.1016/j.avsg.2021.08.050
23. Huang XM, Yu F, Wang Y, et al. Effect of proximal artery restriction on flow reduction and cardiac function in hemodialysis patients with high-flow arteriovenous fistulas. *J Vasc Surg*. Aug 2023;78(2):526-533. doi:10.1016/j.jvs.2023.04.017
24. Janeckova J, Bachleda P, Utikal P, Orsag J. Management of Arteriovenous Fistula After Successful Kidney Transplantation in Long-Term Follow-Up. *Transpl Int*. 2024;37:12841. doi:10.3389/ti.2024.12841
25. Kanno T, Kamijo Y, Hashimoto K, Kanno Y. Outcomes of blood flow suppression methods of treating high flow access in hemodialysis patients with arteriovenous fistula. *Journal of Vascular Access*. November 2015;16(Supplement 10):S28-S33. doi:https://dx.doi.org/10.5301/jva.5000415
26. Kurita N, Mise N, Tanaka S, et al. Arteriovenous Access Closure in Hemodialysis Patients With Refractory Heart Failure: A Single Center Experience. *Therapeutic Apheresis and Dialysis*. April 2011;15(2):195-202. doi:https://dx.doi.org/10.1111/j.1744-9987.2010.00907.x
27. Lee H, Thomas SD, Paravastu S, Barber T, Varcoe RL. Dynamic Banding (DYBAND) Technique for Symptomatic High-Flow Fistulae. *Vascular and Endovascular Surgery*. 01 Jan 2020;54(1):5-11. doi:https://dx.doi.org/10.1177/1538574419874934
28. Miller GA, Goel N, Friedman A, et al. The MILLER banding procedure is an effective method for treating dialysis-associated steal syndrome. *Kidney International*. February 2010;77(4):359-366. doi:https://dx.doi.org/10.1038/ki.2009.461
29. Schier T, Gobel G, Bosmuller C, Gruber I, Tiefenthaler M. Incidence of arteriovenous fistula closure due to high-output cardiac failure in kidney-transplanted patients. *Clinical Transplantation*. November/December 2013;27(6):858-865. doi:https://dx.doi.org/10.1111/ctr.12248
30. Zanow J, Petzold K, Petzold M, Krueger U, Scholz H. Flow reduction in high-flow arteriovenous access using intraoperative flow monitoring. *Journal of Vascular Surgery*. December 2006;44(6):1273-1278. doi:https://dx.doi.org/10.1016/j.jvs.2006.08.010

31. Drouven JW, Wiegersma J, Assa S, et al. Differences in shuntflow (Qa), cardiac function and mortality between hemodialysis patients with a lower-arm fistula, an upper-arm fistula, and an arteriovenous graft. *Journal of Vascular Access*. 2022;doi:https://dx.doi.org/10.1177/11297298221092741
32. Gan W, Zhu F, Mao H, Xiao W, Chen W, Zeng X. Effect of preoperative arterial diameter on hospitalization and mortality in patients undergoing hemodialysis with forearm arteriovenous fistula access. *J Vasc Access*. Nov 23 2023:11297298231211361. doi:10.1177/11297298231211361
33. Hu Z, Wang P, Zeng R, Xu G. Impact of arteriovenous fistula blood flow on serum IL-6, cardiovascular events and death in Chinese patients receiving hemodialysis: A 5-year follow-up. Conference Abstract. *Hong Kong Journal of Nephrology*. October 2015;1):S135. doi:https://dx.doi.org/10.1016/j.hkjn.2015.09.236
34. Quiroga B, Villaverde M, Abad S, Vega A, Reque J, Lopez-Gomez JM. Diastolic dysfunction and high levels of new cardiac biomarkers as risk factors for cardiovascular events and mortality in hemodialysis patients. *Blood Purification*. 24 Apr 2013;36(2):98-106. doi:https://dx.doi.org/10.1159/000354080
35. Repasos E, Kaldara E, Pantsios C, et al. Arteriovenous renal replacement therapy in end-stage left-sided heart failure patients has a detrimental effect on patients with impaired right ventricular function. *Hellenic Journal of Cardiology*. July 2017;58(4):276-280. doi:https://dx.doi.org/10.1016/j.hjc.2016.11.023
36. Wu CK, Wu CL, Lin CH, Leu JG, Kor CT, Tarng DC. Association of vascular access flow with short-term and long-term mortality in chronic haemodialysis patients: A retrospective cohort study. *BMJ Open*. 01 Sep 2017;7(9) (no pagination)e017035. doi:https://dx.doi.org/10.1136/bmjopen-2017-017035
37. Yu Y, Xiong Y, Zhang C, Fu M, Li Y, Fu P. Vascular Access Type Was Not Associated with Mortality and the Predictors for Cardiovascular Death in Elderly Chinese Patients on Hemodialysis. *Blood Purification*. 01 Feb 2020;49(1-2):63-70. doi:https://dx.doi.org/10.1159/000502941
38. Aitken E, Kerr D, Geddes C, Berry C, Kingsmore D. Cardiovascular changes occurring with occlusion of a mature arteriovenous fistula. *J Vasc Access*. Nov-Dec 2015;16(6):459-66. doi:10.5301/jva.5000336
39. Basile C, Vernaglione L, Casucci F, et al. The impact of haemodialysis arteriovenous fistula on haemodynamic parameters of the cardiovascular system. *Clin Kidney J*. Oct 2016;9(5):729-34. doi:10.1093/ckj/sfw063
40. Bos WJW, Zietse R, Wesseling KH, Westerhof N. Effects of arteriovenous fistulas on cardiac oxygen supply and demand. *Kidney International*. 1999;55(5):2049-2053. doi:https://dx.doi.org/10.1046/j.1523-1755.1999.00433.x
41. Chemla ES, Morsy M, Anderson L, Whitemore A. Inflow reduction by distalization of anastomosis treats efficiently high-inflow high-cardiac output vascular access for hemodialysis. *Seminars in Dialysis*. January/February 2007;20(1):68-72. doi:https://dx.doi.org/10.1111/j.1525-139X.2007.00244.x
42. Choi YS, Lee IJ, An JN, et al. High-flow arteriovenous fistula and myocardial fibrosis in hemodialysis patients with non-contrast cardiac magnetic resonance imaging. *Front Cardiovasc Med*. 2022;9:922593. doi:10.3389/fcvm.2022.922593
43. Cridlig J, Selton-Suty C, Alla F, et al. Cardiac impact of the arteriovenous fistula after kidney transplantation: A case-controlled, match-paired study. *Transplant International*. October 2008;21(10):948-954. doi:https://dx.doi.org/10.1111/j.1432-2277.2008.00707.x
44. Dongradi G, Rocha P, Baron B, et al. Hemodynamic effects of arteriovenous fistulae in chronic hemodialysis patients at rest and during exercise. *Clinical Nephrology*. 1981;15(2):75-79.

45. Dundon BK, Torpey DK, Nelson AJ, et al. Beneficial cardiovascular remodeling following arterio-venous fistula ligation post-renal transplantation: a longitudinal magnetic resonance imaging study. *Clin Transplant*. Aug 2014;28(8):916-25. doi:10.1111/ctr.12402
46. Dundon BK, Torpey K, Nelson AJ, et al. The deleterious effects of arteriovenous fistula-creation on the cardiovascular system: A longitudinal magnetic resonance imaging study. *International Journal of Nephrology and Renovascular Disease*. 16 Sep 2014;7:337-345. doi:https://dx.doi.org/10.2147/IJNRD.S66390
47. Elkin DC, Warren JV. Arteriovenous fistulas. Their effect on the circulation. *JAMA (Chicago, Ill)*. 1947;134(18):1524-1528.
48. Hashimoto K, Kanno T, Kamijo Y, Kanno Y. Postoperative maturation and changes in cardiac function differ between types of vascular access. Conference Abstract. *Nephrology Dialysis Transplantation*. June 2020;35(SUPPL 3):iii1675. doi:https://dx.doi.org/10.1093/ndt/gfaa142.P1354
49. Iwashima Y, Horio T, Takami Y, et al. Effects of the creation of arteriovenous fistula for hemodialysis on cardiac function and natriuretic peptide levels in CRF. *American Journal of Kidney Diseases*. 01 Nov 2002;40(5):974-982. doi:https://dx.doi.org/10.1053/ajkd.2002.36329
50. Malik J, Valerianova A, Tuka V, et al. The effect of high-flow arteriovenous fistulas on systemic haemodynamics and brain oxygenation. *ESC Heart Failure*. June 2021;8(3):2165-2171. doi:https://dx.doi.org/10.1002/ehf2.13305
51. Meier M, Stritzke J, Kramer J, et al. Closure of high-volume arteriovenous fistulas improves kidney allograft function in patients with right heart failure. Letter. *Transplant International*. April 2010;23(4):440-443. doi:https://dx.doi.org/10.1111/j.1432-2277.2009.00968.x
52. Ogugua FM, Mathew RO, Ternacle J, Rodin H, Pibarot P, Shroff GR. Impact of arteriovenous fistula on flow states in the evaluation of aortic stenosis among ESKD patients on dialysis. *Echocardiography*. 01 Jan 2024;41(1) (no pagination)e15728. doi:https://dx.doi.org/10.1111/echo.15728
53. Ori Y, Korzets A, Katz M, Perek Y, Zahavi I, Gafter U. Haemodialysis arteriovenous access - A prospective haemodynamic evaluation. *Nephrology Dialysis Transplantation*. January 1996;11(1):94-97. doi:https://dx.doi.org/10.1093/ndt/11.1.94
54. Payne RM, Soderblom RE, Lobstein P. Exercise induced hemodynamic effects of arteriovenous fistulas used for hemodialysis. *Kidney International*. 1972;2(6):344-348. doi:https://dx.doi.org/10.1038/ki.1972.118
55. Polkinghorne KR, Atkins RC, Kerr PG. Determinants of native arteriovenous fistula blood flow. *Nephrology*. August 2004;9(4):205-211. doi:https://dx.doi.org/10.1111/j.1440-1797.2004.00257.x
56. Prastowo RA, Eko Putranto JN, Pratanu I, Intan RE, Alkaff FF. Association between arteriovenous access flow and ventricular function: A cross-sectional study. *Annals of Medicine and Surgery*. May 2022;77 (no pagination)103649. doi:https://dx.doi.org/10.1016/j.amsu.2022.103649
57. Rao NN, Stokes MB, Rajwani A, et al. Effects of Arteriovenous Fistula Ligation on Cardiac Structure and Function in Kidney Transplant Recipients. *Circulation*. 18 Jun 2019;139(25):2809-2818. doi:https://dx.doi.org/10.1161/CIRCULATIONAHA.118.038505
58. Samet P, Bernstein WH, Jacobs W, Fomon J. Indicator-dilution curves in systemic arteriovenous fistulas. *American journal of cardiology*. 1964;13(2):176-187. doi:https://dx.doi.org/10.1016/0002-9149%2864%2990172-9
59. Sandhu JS, Wander GS, Gupta ML, Aulakh BS, Nayyar AK, Sandhu P. Hemodynamic effects of arteriovenous fistula in end-stage renal failure. *Renal Failure*. 2004;26(6):695-701. doi:https://dx.doi.org/10.1081/JDI-200037128
60. Saratzis AN, Saratzis A, Sarafidis PA, Melas N, Ktenidis K, Kiskinis D. Quantitative evaluation of the systemic effects of transposed basilic vein to brachial artery arteriovenous fistula: A prospective study. *Journal of Vascular Access*. 2008;9(4):285-290. doi:https://dx.doi.org/10.1177/112972980800900410

61. Saritas B, Okyay K, Yilmazturk H. Perforating vein-brachial artery anastomosis as an alternative to the conventional arterio-venous fistulae for haemodialysis: Mid-term follow-up results. *Turkiye Klinikleri Cardiovascular Sciences*. 2010;22(2):200-205.
62. Sheashaa H, Hassan N, Osman Y, Sabry A, Sobh M. Effect of spontaneous closure of arteriovenous fistula access on cardiac structure and function in renal transplant patients. *American Journal of Nephrology*. 2004;24(4):432-437. doi:https://dx.doi.org/10.1159/000080187
63. Tang CY, Zhu CP, Wang RP, et al. Effect of Blood Pump Flow and Arteriovenous Fistula Blood Flow on the Blood Pressure and Cardiac Function in Patients Undergoing Maintenance Hemodialysis. *Therapeutic Apheresis and Dialysis*. 01 Dec 2019;23(6):556-561. doi:https://dx.doi.org/10.1111/1744-9987.12809
64. Tellioglu G, Berber I, Kilicoglu G, Seymen P, Kara M, Titiz I. Doppler Ultrasonography-Guided Surgery for High-Flow Hemodialysis Vascular Access: Preliminary Results. *Transplantation Proceedings*. January 2008/February 2008;40(1):87-89. doi:https://dx.doi.org/10.1016/j.transproceed.2007.11.059
65. Unger P, Xhaet O, Wissing KM, Najem B, Dehon P, van de Borne P. Arteriovenous fistula closure after renal transplantation: a prospective study with 24-hour ambulatory blood pressure monitoring. *Transplantation*. 15 Feb 2008;85(3):482-485.
66. Valek M, Lopot F, Polakovic V. Arteriovenous fistula, blood flow, cardiac output, and left ventricle load in hemodialysis patients. *ASAIO journal (American Society for Artificial Internal Organs : 1992)*. 2010 2010;56(3):200-203.
67. Velez-Roa S, Neubauer J, Wissing M, et al. Acute arterio-venous fistula occlusion decreases sympathetic activity and improves baroreflex control in kidney transplanted patients. *Nephrology Dialysis Transplantation*. June 2004;19(6):1606-1612. doi:https://dx.doi.org/10.1093/ndt/gfh124
68. Warren JV, Brannon ES, Cooper Jr FW. The hemodynamics of rapid changes in cardiac output in man. *Journal of Clinical Investigation*. 1947;26(6):1199.
69. Warren JV, Nickerson JI, Elkin DC. I. The cardiac output in patients with arteriovenous fistulas. II. The effect of temporary occlusion of arteriovenous fistulas on heart rate, stroke volume and cardiac output. III. The blood volume in patients with arteriovenous fistulas. *Journal of Clinical Investigation*. 1951;30(2):210-226.
70. Wijnen E, Keuter XH, Planken NR, et al. The relation between vascular access flow and different types of vascular access with systemic hemodynamics in hemodialysis patients. *Artificial Organs*. December 2005;29(12):960-964. doi:https://dx.doi.org/10.1111/j.1525-1594.2005.00165.x
71. Ye WL, Fang LG, Ma J, Li XM. Long-term effects of arteriovenous fistula on cardiac structure and function in non-diabetic hemodialysis patients. [Chinese]. *Acta Academiae Medicinae Sinicae*. 28 Feb 2013;35(1):95-101. doi:https://dx.doi.org/10.3881/j.issn.1000-503X.2013.01.018
72. Glowinski J, Malyszko J, Glowinska I, Mysliwiec M. To close or not to close: Fistula ligation and cardiac function in kidney allograft recipients. *Polskie Archiwum Medycyny Wewnetrznej*. 2012;122(7-8):348-352. doi:https://dx.doi.org/10.20452/pamw.1349
73. Hiremath S, Doucette SP, Richardson R, Chan K, Burns K, Zimmerman D. Left ventricular growth after 1 year of haemodialysis does not correlate with arteriovenous access flow: A prospective cohort study. *Nephrology Dialysis Transplantation*. August 2010;25(8):2656-2661. doi:https://dx.doi.org/10.1093/ndt/gfq081
74. He X, Liu Y. Effects of arteriovenous fistulas and central venous catheters on the cardiac function and prognosis of patients on maintenance hemodialysis. *Pak J Med Sci*. May-Jun 2023;39(3):780-784. doi:10.12669/pjms.39.3.7151

75. Jaques DA, Davenport A. High-flow arteriovenous fistula is not associated with increased extracellular volume or right ventricular dysfunction in haemodialysis patients. *Nephrology Dialysis Transplantation*. 01 Mar 2021;36(3):536-543. doi:https://dx.doi.org/10.1093/ndt/gfaa188
76. Laranjinha I, Matias P, Azevedo A, et al. Are high flow arteriovenous accesses associated with worse haemodialysis? *Jornal brasileiro de nefrologia : 'orgao oficial de Sociedades Brasileira e Latino-Americana de Nefrologia*. 01 Apr 2018;40(2):136-142. doi:https://dx.doi.org/10.1590/2175-8239-JBN-3875
77. Movilli E, Viola BF, Brunori G, et al. Long-term Effects of Arteriovenous Fistula Closure on Echocardiographic Functional and Structural Findings in Hemodialysis Patients: A Prospective Study. *American Journal of Kidney Diseases*. April 2010;55(4):682-689. doi:https://dx.doi.org/10.1053/j.ajkd.2009.11.008
78. Saleh MA, El Kilany WM, Keddis VW, El Said TW. Effect of high flow arteriovenous fistula on cardiac function in hemodialysis patients. *Egyptian Heart Journal*. December 2018;70(4):337-341. doi:https://dx.doi.org/10.1016/j.ehj.2018.10.007
79. Salehi T, Montarello NJ, Juneja N, et al. Long-Term Impact of Arteriovenous Fistula Ligation on Cardiac Structure and Function in Kidney Transplant Recipients: A 5-Year Follow-Up Observational Cohort Study. *Kidney360*. Jul 29 2021;2(7):1141-1147. doi:10.34067/kid.0000692021
80. Song W, Wu L, Sun C, Kong X, Wang H. New-onset atrial fibrillation following arteriovenous fistula increases adverse clinical events in dialysis patients with end-stage renal disease. *Front Cardiovasc Med*. 2024;11:1386304. doi:10.3389/fcvm.2024.1386304
81. Unger P, Wissing KM, de Pauw L, Neubauer J, van de Borne P. Reduction of left ventricular diameter and mass after surgical arteriovenous fistula closure in renal transplant recipients. Clinical Trial; Controlled Clinical Trial; Journal Article; Research Support, Non-U.S. Gov't. *Transplantation*. 2002;74(1):73-79. doi:10.1097/00007890-200207150-00013
82. Valerianova A, Malik J, Janeckova J, et al. Reduction of arteriovenous access blood flow leads to biventricular unloading in haemodialysis patients. *International Journal of Cardiology*. 01 Jul 2021;334:148-153. doi:https://dx.doi.org/10.1016/j.ijcard.2021.04.027
83. Van Duijnhoven ECM, Cheriex ECM, Tordoir JHM, Kooman JP, Van Hooff JP. Effect of closure of the arteriovenous fistula on left ventricular dimensions in renal transplant patients. *Nephrology Dialysis Transplantation*. 2001;16(2):368-372. doi:http://dx.doi.org/10.1093/ndt/16.2.368
84. Wohlfahrt P, Rokosny S, Melenovsky V, Borlaug BA, Pecenkova V, Balaz P. Cardiac remodeling after reduction of high-flow arteriovenous fistulas in end-stage renal disease. *Hypertension Research*. 01 Sep 2016;39(9):654-659. doi:https://dx.doi.org/10.1038/hr.2016.50
85. Beigi AA, Sadeghi AMM, Khosravi AR, Karami M, Masoudpour H. Effects of the arteriovenous fistula on pulmonary artery pressure and cardiac output in patients with chronic renal failure. *Journal of Vascular Access*. 2009;10(3):160-166. doi:https://dx.doi.org/10.1177/112972980901000305
86. Chen R, Wu X, Shen LJ, et al. Left ventricular myocardial function in hemodialysis and nondialysis uremia patients: A three-dimensional speckle-tracking echocardiography study. *PLoS ONE*. 24 Jun 2014;9(6) (no pagination)e100265. doi:https://dx.doi.org/10.1371/journal.pone.0100265
87. De Lima JJG, Campos Vieira ML, Molnar LJ, Medeiros CJ, Ianhez LE, Krieger EM. Cardiac effects of persistent hemodialysis arteriovenous access in recipients of renal allograft. *Cardiology*. 1999;92(4):236-239. doi:http://dx.doi.org/10.1159/000006980
88. Di Lullo L, Floccari F, Polito P. Right ventricular diastolic function in dialysis patients could be affected by vascular access. *Nephron - Clinical Practice*. June 2011;118(3):c257-c261. doi:https://dx.doi.org/10.1159/000321867

89. Paneni F, Gregori M, Ciavarella GM, et al. Relation between right and left ventricular function in patients undergoing chronic dialysis. *Journal of Cardiovascular Medicine*. April 2013;14(4):289-295. doi:https://dx.doi.org/10.2459/JCM.0b013e32834eacf0
90. Papasotiriou M, Xanthopoulou I, Ntrinias T, et al. Impact of arteriovenous fistula on cardiac size and function in kidney transplant recipients: A retrospective evaluation of 5-year echocardiographic outcome. *Experimental and Clinical Transplantation*. 2019;17(5):619-626. doi:https://dx.doi.org/10.6002/ect.2018.0331
91. Said K, Hassan M, Farouk M, Baligh E, Zayed B. Right Ventricular Function After Creation of an Atriovenous Fistula in Patients With End Stage Renal Disease. *Heart Lung Circ*. Jun 2019;28(6):884-892. doi:10.1016/j.hlc.2018.04.282
92. Tayebi P, Ziaie N, Golshan S, Bijani A, Mahmoudlou F. Hemodialysis Patients with High-Flow Arteriovenous Fistulas: An Evaluation of the Impact on Cardiac Function. *Vasc Specialist Int*. Mar 8 2024;40:7. doi:10.5758/vsi.230090
93. Abdelwhab S, Elshinnawy S. Pulmonary hypertension in chronic renal failure patients. *Am J Nephrol*. 2008;28(6):990-7. doi:10.1159/000146076
94. Ori Y, Korzets A, Katz M, et al. The contribution of an arteriovenous access for hemodialysis to left ventricular hypertrophy. *American Journal of Kidney Diseases*. 01 Oct 2002;40(4):745-752. doi:https://dx.doi.org/10.1053/ajkd.2002.35685
95. Abdelghany MF, Mohamad WH, Elden AB. Pulmonary hypertension in patients with end-stage renal disease under regular hemodialysis: A cross-sectional study. *Egyptian Journal of Chest Diseases and Tuberculosis*. January-March 2020;69(1):235-241. doi:https://dx.doi.org/10.4103/ejcdt.ejcdt_38_19
96. Acarturk G, Albayrak R, Melek M, et al. The relationship between arteriovenous fistula blood flow rate and pulmonary artery pressure in hemodialysis patients. *International urology and nephrology*. 2008;40(2):509-513. doi:https://dx.doi.org/10.1007/s11255-007-9269-8
97. Ayyaz A, Khan AA, Akram M, Ahmed F, Asif M, Hameed F. To assess the correlation of arteriovenous fistula flow with pulmonary hypertension in end stage renal disease. *Pakistan Journal of Medical and Health Sciences*. October 2020;14(4):928-930.
98. Emara MM, Habeb MA, Alnahal AA, Elshazly TA, Alatawi FO, Masoud AS. Prevalence of pulmonary hypertension in patients with chronic kidney disease on and without dialysis. *Egyptian Journal of Chest Diseases and Tuberculosis*. October 2013;62(4):761-768. doi:https://dx.doi.org/10.1016/j.ejcdt.2013.09.011
99. Havlucu Y, Kursat S, Ekmekci C, et al. Pulmonary hypertension in patients with chronic renal failure. *Respiration*. August 2007;74(5):503-510. doi:https://dx.doi.org/10.1159/000102953
100. Reque J, Garcia-Prieto A, Linares T, et al. Pulmonary Hypertension Is Associated with Mortality and Cardiovascular Events in Chronic Kidney Disease Patients. *American Journal of Nephrology*. 01 Feb 2017;45(2):107-114. doi:https://dx.doi.org/10.1159/000453047
101. Tudoran M, Ciocarlie T, Mates A, Pescariu SA, AbuAwwad A, Tudoran C. Pulmonary hypertension in patients with end stage renal disease undergoing hemodialysis. *Niger J Clin Pract*. Feb 2020;23(2):198-204. doi:10.4103/njcp.njcp_278_19
102. Warner ED, Corsi DR, Jimenez D, et al. Determinants of pulmonary hypertension in patients with end-stage kidney disease and arteriovenous access. *Curr Probl Cardiol*. Apr 2024;49(4):102406. doi:10.1016/j.cpcardiol.2024.102406
103. Yigla M, Nakhoul F, Sabag A, et al. Pulmonary hypertension in patients with end-stage renal disease. *Chest*. 01 May 2003;123(5):1577-1582. doi:https://dx.doi.org/10.1378/chest.123.5.1577
104. Zhao LH, S.; Liang, T; Tang, H. Effects of maintenance hemodialysis on right ventricular dysfunction in patients with end-stage renal disease. [Chinese]. *Sichuan da xue xue bao*. 01 Sep 2014;Yi xue ban = Journal of Sichuan University. Medical science edition. 45(5):814-818.

105. Adachi T, Sakurada T, Otowa T, et al. Impact of vascular access intervention therapy on cardiac load in hemodialysis patients. *Hemodial Int*. Oct 2016;20 Suppl 1:S12-s16. doi:10.1111/hdi.12460
106. Malik J, Tuka V, Krupickova Z, Chytilova E, Holaj R, Slavikova M. Creation of dialysis vascular access with normal flow increases brain natriuretic peptide levels. *International Urology and Nephrology*. 2009;41(4):997-1002. doi:https://dx.doi.org/10.1007/s11255-009-9544-y
107. Stoumpos S, Rankin A, Hall Barrientos P, et al. Interrogating the haemodynamic effects of haemodialysis arteriovenous fistula on cardiac structure and function. *Scientific reports*. 13 Sep 2021;11(1):18102. doi:https://dx.doi.org/10.1038/s41598-021-97625-5
